# Stay-at-Home Orders, African American Population, Poverty and State-level Covid-19 Infections: Are there associations?

**DOI:** 10.1101/2020.06.17.20133355

**Authors:** Sangeetha Padalabalanarayanan, Vidya Sagar Hanumanthu, Bisakha P. Sen

**Affiliations:** Department of Health Services Administration, University of Alabama at Birmingham, Birmingham, AL, USA; School of Health Professions, University of Alabama at Birmingham, Birmingham, AL, USA; Division of Clinical Immunology and Rheumatology, University of Alabama at Birmingham, Birmingham, Alabama, USA; Department of Health Care Organization and Policy, University of Alabama at Birmingham, Birmingham, AL, USA

## Abstract

**Importance:** To cope with the continuing COVID-19 pandemic, state and local health officials need information on the effectiveness of policies aimed at curbing contagion, as well as area-specific socio-demographic characteristics that can portend vulnerability to the disease.

**Objective:** To investigate whether state-imposed stay-at-home orders, African American population in the state, state poverty and other state socio-demographic characteristics, were associated with the state-level incidence of COVID-19 infection.

**Design, Setting, Participants:** State-level, aggregated, publicly available data on positive COVID-19 cases and tests were used. The period considered was March 1st-May 4th. All U.S. states except Washington were included. Outcomes of interest were daily cumulative and daily incremental COVID-19 infection rates. Outcomes were log-transformed. Log-linear regression models with a quadratic time-trend and random intercepts for states were estimated. Covariates included log-transformed test-rates, a binary indicator for stay-at-home, percentage of African American, poverty, percentage elderly, state population and prevalence of selected comorbidities. Binary ‘fixed effects’ for date each state first started reporting test data were included.

**Results:** Stay-at-home orders were associated with decreases in cumulative (β:-1.23; T-stat: - 6.84) and daily (β:-0.46; T-stat: −2.56) infection-rates. Predictive analyses indicated that expected cumulative infection rates would be 3 times higher and expected daily incremental rates about 60% higher in absence of stay-at-home orders. Higher African American population was associated with higher cumulative (β: 0.08; T-stat: 4.01) and daily (β: 0.06; T-stat: 3.50) rates. State poverty rates had mixed results and were not robust to model specifications. There was strong evidence of a quadratic daily trend for cumulative and daily rates. Results were largely robust to alternate specifications.

**Conclusions:** We find evidence that stay-at-home orders, which were widely supported by public-health experts, helped to substantially curb COVID-19 infection-rates. As we move to a phased re-opening, continued precautions advised by public-health experts should be adhered to. Also, a larger African American population is strongly associated with incidence of COVID-19 infection. Policies and resources to help mitigate African American vulnerability to COVID-19 is an urgent public health and social justice issue, especially since the ongoing mass protests against police brutality may further exacerbate COVID-19 contagion in this community.

**Key Points:** *Question:* Did the stay-at-home orders, African American population and other socio demographic factors across states have any associations with COVID-19 infection rates across states?

*Findings:* Multivariate log-linear regression models using daily state level data from March-May found evidence that when stay-at-home orders were implemented, they helped reduce state COVID-19 cumulative and daily infection rates substantially. Further, we found that states with larger African-American population had higher COVID-19 infection rates.

*Meaning:* Results suggest that state-level stay-at-home orders helped reduce COVID-19 infection rates substantially, and also that African American populations may be especially vulnerable to COVID-19 infection.

## Introduction

COVID-19 is cause by novel coronavirus, SARS-CoV-2, that mainly spreads through person to person contact. This coronavirus has spread rapidly across countries, and on March 12^th^ World Health Organization declared COVID-19 as a pandemic^1^. One epidemiological model predicted that in absence of interventions or mitigation strategies, worldwide deaths may reach 40 million^2^. COVID-19 led to unprecedented shutdowns worldwide in an effort to curb the contagion. Currently, however, countries, including the US, are moving into phased reopening, which bring with it a different set of uncertainties about associated health-risks^3^.

Our understanding of risk-factors for COVID-19 infection and outcomes are still at fairly early stages, including the effectiveness of the shutdown. A bivariate analysis published in Wall Street Journal argued that shutdowns did not curb COVID-19 fatalities^4^. However, more recent analyses using data from selected U.S. counties or states has suggested shutdowns helped curb infections, hospitalizations and deaths^5,6^.^7^ Mixed messages from political authorities in U.S. on the usefulness of shutdowns has often conflicted with advice offered by public-health scientists.^8^ The resulting cynicism may undermine the effectiveness of public-health advice and messaging on continued safety measures after re-opening, like using face-masks, social distancing, and diligent hand-hygiene^8^.

Our understanding of the role of socio-demographic risk-factors is similarly incomplete. There is indication that infection rates are higher among African-American communities based on reports from selected areas of the country^9^. One analyses in New York City showed that neighborhoods with higher concentrations of minorities and low-income populations had higher infection rates^10^. Given the highly contagious nature of disease, there is speculation that population density may increase infection rates. Also, while being elderly or having pre-existing conditions like diabetes or cardiovascular disease are linked to adverse health consequences after infection^9^, it is unclear whether there are associations between these factors and the risk of infection per se.

We present an ecological study using state-level data to quantitatively analyze risk-factors for state-level COVID-19 incidence. Specifically, we investigated associations between state-mandated shutdowns, state socio-demographic characteristics – especially African American population and poverty -- and reported COVID-19 infection rates. We use the terms ‘stay-at-home’ and ‘shutdown’ interchangeably, since the literature does not clearly differentiate between the two. Our findings add to the small but growing body of literature on stay-at-home orders, ecological risk-factors and COVID-19 which can help inform public-health policymakers.

## Methods

We conducted multivariate regression analyses of state-level predictors on the incidence of COVID-19 using state-level observational data. Data on COVID-19 positive cases (hereafter ‘cases’) and COVID-19 tests (hereafter ‘tests’) are obtained from ‘The COVID Tracking Project’ (TCTP)^11^. Initiated by *The Atlantic* in partnership with *Related Sciences*, TCTP is a collaboration of multiple stakeholders and volunteers, who collate data from state health agencies and make it publicly available to researchers and journalists. One unique advantage of TCTP is that they include daily data on tests as well as cases, and their testing data is used by other disseminators of COVID-19 information such as 1point3acres^12^.

We used daily case and test data over March 1^st^-May 4^th^, 2020. Prior to March 1^st^, only data from the state of Washington was available, where the first case was detected on January 22^nd^. Other states started reporting between March 1 and March 12. We stopped at May 4^th^, 2020, since states started to lift stay-at-home restrictions from early May. Figure.1 shows the distribution of daily cumulative cases and rates by state. Figure. 2 shows the trend in cumulative rates by state. Reporting start-date for each state and other state-specific pertinent information are shown in supplement Table 1.

**Figure.1a.**
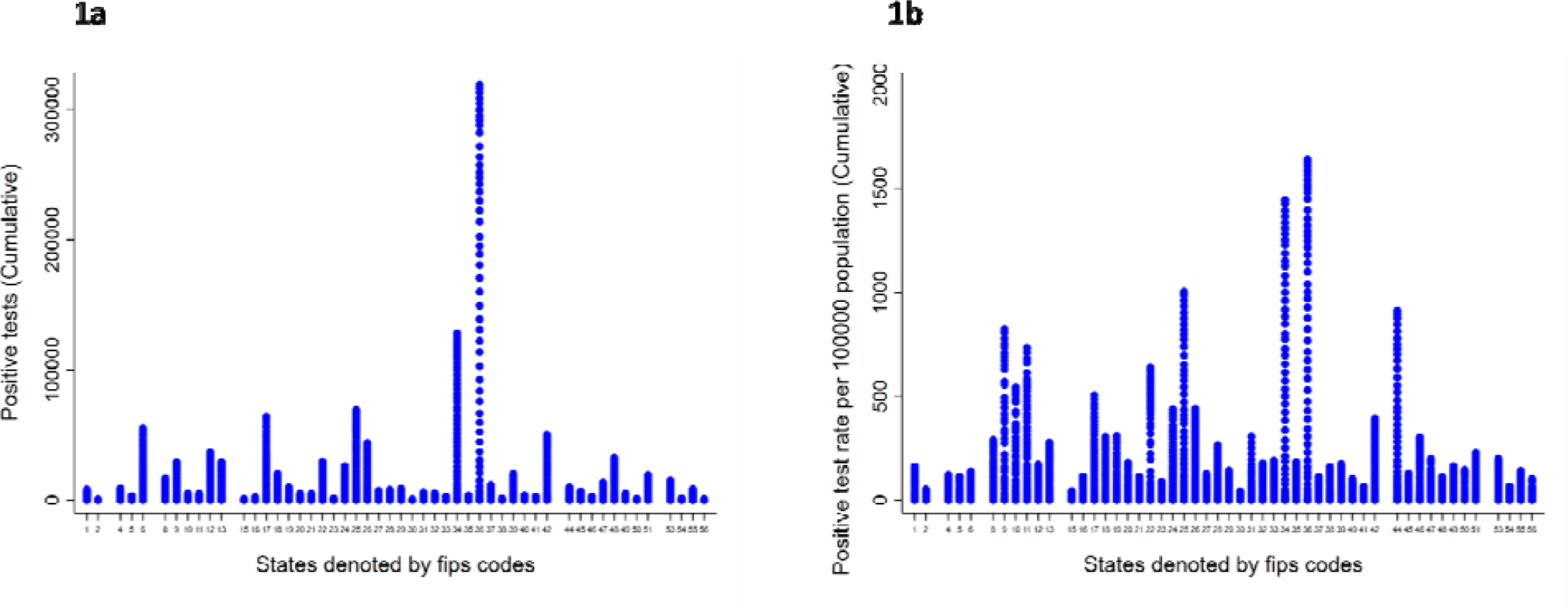
Cumulative positive test counts by states. 1b. Cumulative positive test rate by state.

**Figure.2.**
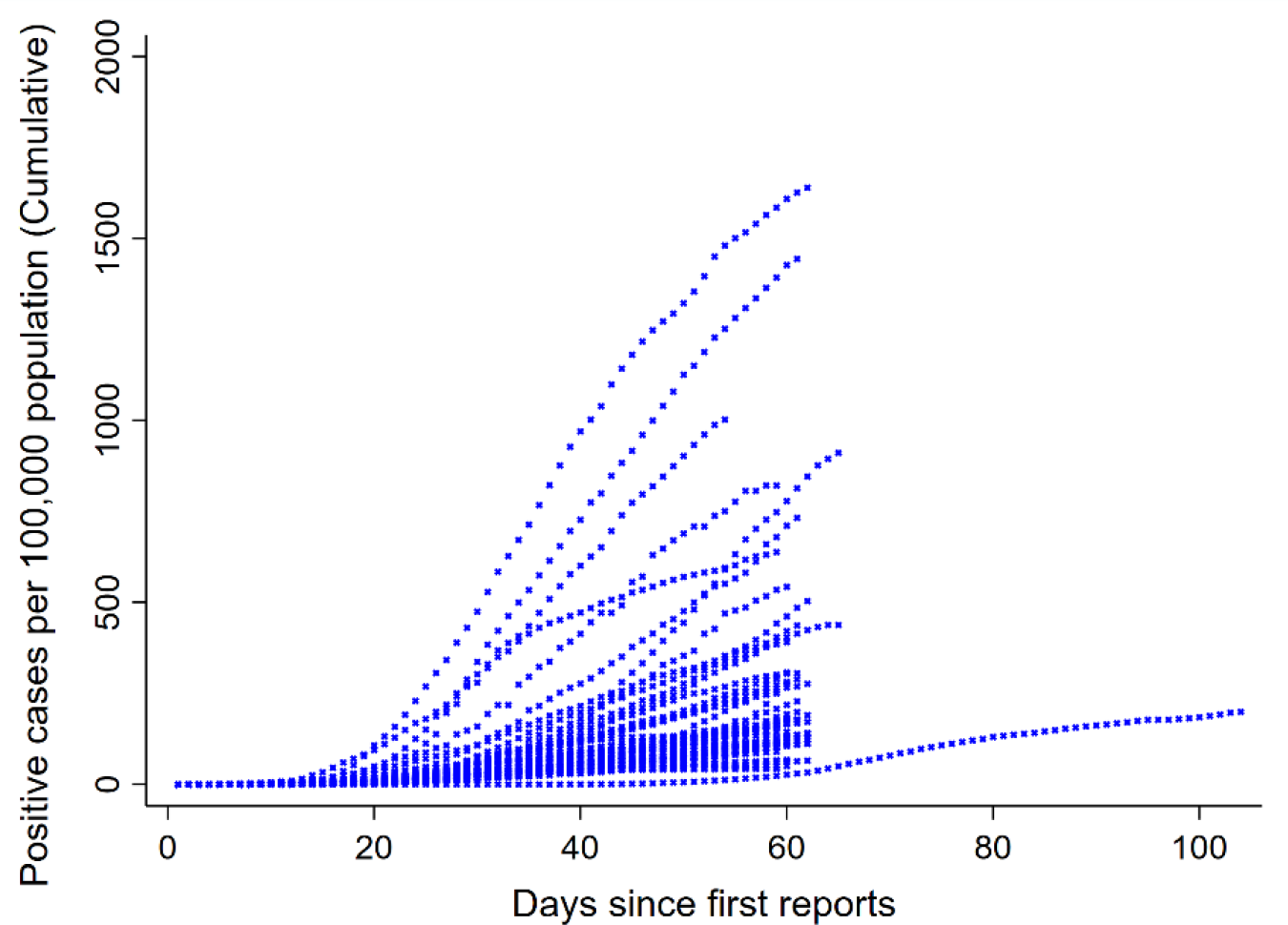
Cumulative positive tests reported by state.

### Outcome variable

Our outcome variable of interest was cases per 100,000 population by state and day. We examined both daily cumulative cases, and daily incremental cases. These were log-transformed to de-emphasize the impact of outliers, such as the state of New York.

### Independent variables

All models controlled for cumulative and daily tests per 100,000 population, and included a quadratic time-trend. Our main independent variables of interest were the percentage of state population who are African-American^13^, and a binary indicator for whether a stay-at-home order has been issued in the state versus not^14-16^. Other control variables included both total population^17^ and population density^18^, state poverty rate ^19^, the percentage of population above 65 years^20^ and the prevalence of asthma and diabetes in the state^20^. As part of our sensitivity analyses, we also included physicians per 100,000 state population^21^ and certified nursing facilities per 100,000 population^22^ in the model.

We estimated log-linear multivariate regression models, with quadratic time trends and state random intercepts, in the following form:

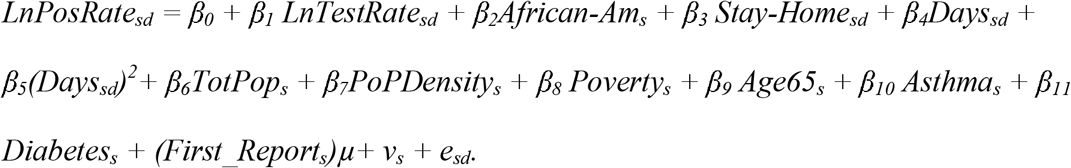

Models were estimated using daily cumulative cases and tests and daily incremental cases and tests for each state. A quadratic specification for days (Days_sd_ and Days_sd_^2^) was included to accommodate for non-linear daily trends. Stay-Home_sd_ was a binary indicator with 1 if stay-at-home orders had been issued in the state and 0 if not. First_Report_s_ is a vector of dummy variables or ‘fixed effects’ for the date when a state’s test and case data first became available in TCTP. This is included since variations in date of first report may be indicative of variations in the seriousness of the outbreak in a state, as well as awareness and attitudes among state authorities to COVID-19 risk, which can be correlated issue of stay-at-home orders. The other covariates are time-invariant for our study period. v_s_, the random intercept, captures the normally distributed heterogeneity between states.

Because the outcome variable is logarithmically transformed, coefficient estimates can be interpreted as percentage changes in the outcome variable using the formula 100[exp(β_j_) −1]. To obtain predicted values of the outcome variables under different scenarios, we use the retransformation method recommended by Cameron &Trivedi ^23^. We used STATA version 16 (College Station, Texas, USA) for all empirical analyses, and ‘xtreg, re’ command to estimate our model.

## Results

Our dataset has 3127 day-level observations where cumulative positive cases and cumulative tests were reported. Daily incremental cases and tests are available for 3113 and 3105 observations. The missing values are because the incremental tests and cases were not reported for the very first day the data for some states were available. Pooled summary statistics for outcome and control variables of interest are summarized in Table 1. Mean cumulative positive rates (calculated per 100,000 state population) cases is 101.6, and mean test rate is 744.82. Mean daily case rates is 5.08 per 100,000 population, and mean daily test rates is 38.63 per 100,000.

**Table.1.** Descriptive Statistics

| **Table.1. Descriptive Statistics** |  |  |  |
| --- | --- | --- | --- |
| **Variable** | **Observations** | **Mean** | **Std.Dev.** |
| Daily cumulative positive case rate<sup>a</sup> | 3127 | 101.62 | 197.31 |
| Daily cumulative test rate | 3127 | 744.82 | 895.50 |
| Daily incremental positive case rate | 3113 | 5.08 | 8.37 |
| Daily incremental test rate | 3105 | 38.63 | 50.65 |
| Log cumulative positive case rate | 3127 | 2.02 | 5.03 |
| Log cumulative test rate | 3127 | 4.76 | 3.57 |
| Log incremental positive case rate | 3111 | -1.14 | 5.51 |
| Log incremental test rate | 3098 | 15.57 | 8.75 |
| Stay-at-home in place (1 if in place, 0 otherwise) | 3127 | 0.54 | 0.50 |
| Population Density<sup>b</sup> | 3127 | 0.04 | 0.16 |
| Total Population<sup>c</sup> | 3127 | 65.22 | 73.06 |
| Asthma<sup>d</sup> | 3127 | 9.68 | 1.21 |
| Diabetes<sup>e</sup> | 3127 | 11.10 | 1.89 |
| African American<sup>f</sup> | 3127 | 10.84 | 10.38 |
| Above-65<sup>g</sup> | 3127 | 16.46 | 2.04 |
| Poverty<sup>h</sup> | 3127 | 11.76 | 2.80 |

<sup>a</sup>Rate= per 100000 of population; <sup>b</sup>Population Density:10000 persons per Square mile (Year 2020); <sup>c</sup>Total Population per 100000 (2019) <sup>d</sup>Asthma: Crude percentage of population with Asthma per state (2018); <sup>e</sup>Diabetes: Crude percentage of population with diabetes per state (2018); <sup>f</sup>African American: African American population percentage per state (2018); <sup>g</sup>Above-65: Percentage of population above 65 years of age in a state (2018); <sup>h</sup>Poverty: Percentage of population under poverty per state (2017-2018).

Stay-at-home orders were in place during 54 percent of days in our sample. The percent of African-Americans in the pooled state-day sample was 10.85, the percent in poverty was 11.76, and prevalence of asthma and diabetes were respectively 9.68 percent and 11.10 percent. Detailed state by state data is reported in supplemental Table I.

We examined the trends in the positive test cumulative (Figure.1a) and rate (Figure.1b) among states. We observed a disproportionate number of cases reported in states such as New York and New Jersey. To offset effects of these states, that are apparent outliers, we log-transformed the case rates as well as test rates. When log-transforming, ‘zero’ was replaced by 1/10million. For 2 observations in daily incremental cases and 7 observations for daily incremental tests, the numbers were negative – representing a downward revision of numbers reported the previous day. Those observations were dropped prior to log transformation.

Daily trends in reported cases indicate a quadratic trend that increases at a decreasing rate (Figure.2). The state of Washington started reporting COVID-19 data earlier than any other states because that state had one confirmed case on January 20. There was also sporadic testing in February in WA that government officials later shut down^24^. Due to these distinctive experiences of WA, we did not include that state in our main regression analyses. We tested the sensitivity of our results to excluding New York (the state with highest cases), to including Washington, and to omitting state-level observations where states had not yet started reporting any tests. The results are in Table 2 and supplemental Table II.

**Table.2.**
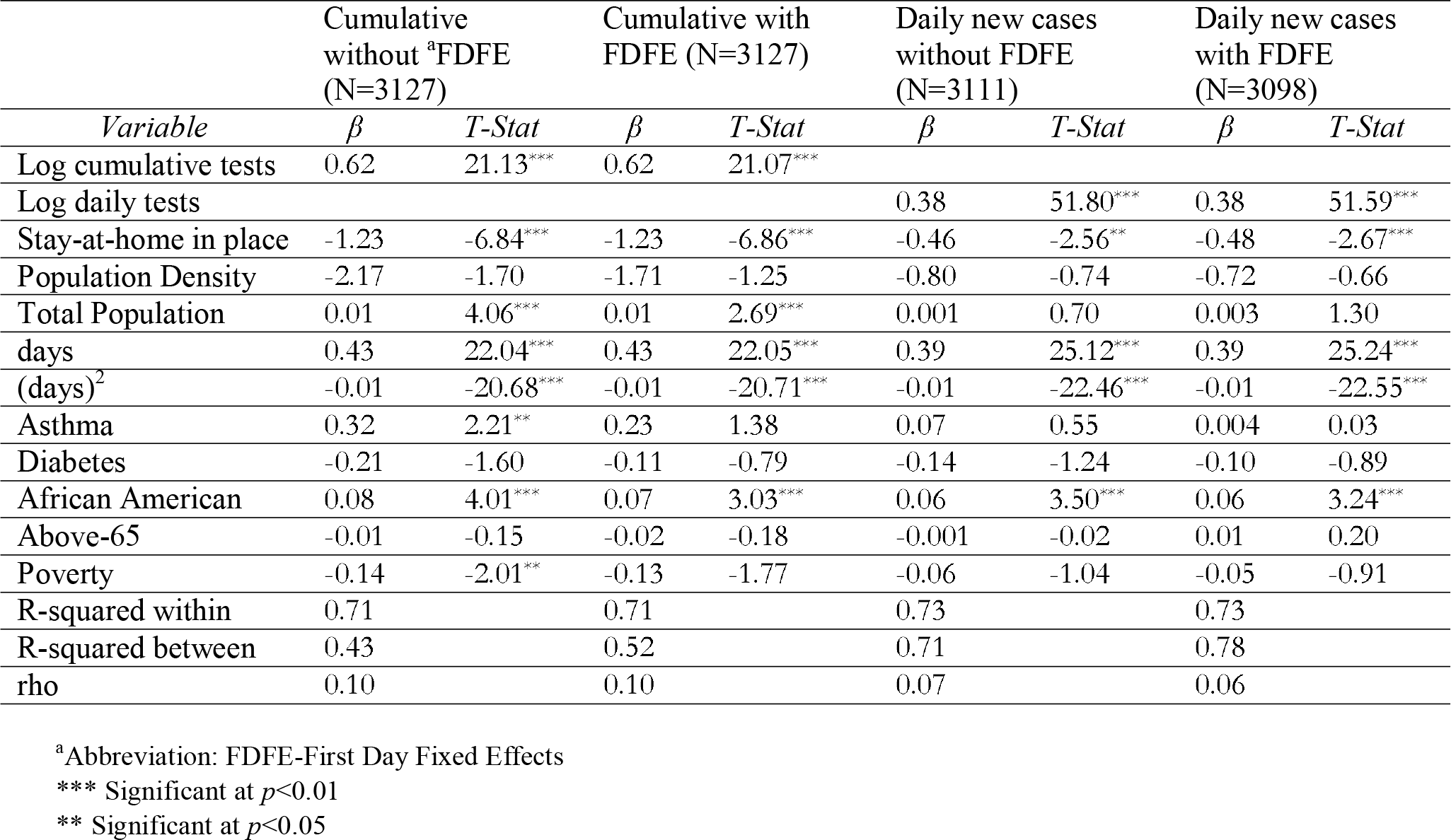
Regression Analysis with State Random Effects, for Cumulative and Daily COVID-19 Cases Main Model.

| **Table.2. Regression Analysis with State Random Effects, for Cumulative and Daily COVID-19 Cases** |  |  |  |  |  |  |  |  |
| --- | --- | --- | --- | --- | --- | --- | --- | --- |
| **Main Model** |  |  |  |  |  |  |  |  |
|  | Cumulative without <sup>a</sup>FDFE (N=3127) |  | Cumulative with FDFE (N=3127) |  | Daily new cases without FDFE (N=3111) |  | Daily new cases with FDFE (N=3098) |  |
| *Variable* | $\beta$ | *T-Stat* | $\beta$ | *T-Stat* | $\beta$ | *T-Stat* | $\beta$ | *T-Stat* |
| Log cumulative tests | 0.62 | 21.13*** | 0.62 | 21.07*** |  |  |  |  |
| Log daily tests |  |  |  |  | 0.38 | 51.80*** | 0.38 | 51.59*** |
| Stay-at-home in place | -1.23 | -6.84*** | -1.23 | -6.86*** | -0.46 | -2.56** | -0.48 | -2.67*** |
| Population Density | -2.17 | -1.70 | -1.71 | -1.25 | -0.80 | -0.74 | -0.72 | -0.66 |
| Total Population | 0.01 | 4.06*** | 0.01 | 2.69*** | 0.001 | 0.70 | 0.003 | 1.30 |
| days | 0.43 | 22.04*** | 0.43 | 22.05*** | 0.39 | 25.12*** | 0.39 | 25.24*** |
| (days)<sup>2</sup> | -0.01 | -20.68*** | -0.01 | -20.71*** | -0.01 | -22.46*** | -0.01 | -22.55*** |
| Asthma | 0.32 | 2.21** | 0.23 | 1.38 | 0.07 | 0.55 | 0.004 | 0.03 |
| Diabetes | -0.21 | -1.60 | -0.11 | -0.79 | -0.14 | -1.24 | -0.10 | -0.89 |
| African American | 0.08 | 4.01*** | 0.07 | 3.03*** | 0.06 | 3.50*** | 0.06 | 3.24*** |
| Above-65 | -0.01 | -0.15 | -0.02 | -0.18 | -0.001 | -0.02 | 0.01 | 0.20 |
| Poverty | -0.14 | -2.01** | -0.13 | -1.77 | -0.06 | -1.04 | -0.05 | -0.91 |
| R-squared within | 0.71 |  | 0.71 |  | 0.73 |  | 0.73 |  |
| R-squared between | 0.43 |  | 0.52 |  | 0.71 |  | 0.78 |  |
| rho | 0.10 |  | 0.10 |  | 0.07 |  | 0.06 |  |

<sup>a</sup>Abbreviation: FDFE-First Day Fixed Effects
\*\*\* Significant at $p < 0.01$
\*\* Significant at $p < 0.05$

Results from our main model are in Table 2. Models are estimated both with and without ‘First Day Fixed Effects’ (FDFE), a vector of binary variables capturing the first-date of reporting for each state. For cumulative case rates, stay-at-home orders were significantly associated with a reduction in cases (β: −1.23, T-stat: −6.84), and this remained essentially unchanged with FDFE (β: −1.23, T-stat: −6.86). Applying 100[exp(β_j_) −1] to interpret results as percentage changes, this implies that in the absence of stay-at-home orders, cumulative case rates would on average be approximately 240 percent higher. Results remain robust to other sensitivity analyses like excluding state of New York (β: −1.22, T-stat: −6.71 without FDFE, β: −1.23, T-stat: −6.74 with FDFE; supplemental Table IIA), and dropping the state-day observations when no cumulative tests had yet been reported (β:-1.23 T-stat: −6.79 without FDFE) in supplement Table IIC. When observations for WA were added to the model, results continued to be negative and significant, albeit smaller in magnitude, without and with FDFE (β: −0.61, T-stat: −3.51; β: −0.60, T-stat: −3.42 respectively; supplemental Table IIB).

On examining the daily new cases, we observed the same pattern for stay-at-home orders for both without (β: −0.46, T-stat: −2.56) and with FDFE (β: −0.48, T-stat: −2.67). Interpreting these as percentage changes imply that, in the absence of stay-at-home orders, daily cases would, on average, be almost 60 percent higher than with the orders. These results hold for sensitivity analyses, with the exception of the models where Washington is.

In Table 3, we show the expected predicted values of cumulative positive case rates and daily positive case rates with no stay-at-home orders (stay-at-home=0) versus stay-at-home orders (stay-at-home=1) at different intervals in time, holding all other variables in the model at the mean values. This confirms that expected cumulative cases are 2.5-3 times higher and expected daily cases are about 60% higher in the absence of stay-at-home.

**Table.3.** Predicted Rates With & Without Stay-at-Home

| **Table.3. Predicted Rates With & Without Stay-at-Home** |  |  |  |  |
| --- | --- | --- | --- | --- |
| Days since first reporting | Expected Predicted Value of Cumulative Case Rate with Stay-at-home=0 | Expected Predicted Value of Cumulative Case Rate with Stay-at-home=1 | Expected Predicted Value of Daily Case Rate with Stay-at-home=0 | Expected Predicted Value of Daily Case Rate with Stay-at-home=1 |
| Day 05 | 13.5 | 4.0 | 0.83 | 0.53 |
| Day 10 | 78.7 | 23.1 | 4.0 | 2.5 |
| Day 15 | 358.4 | 105.2 | 15.5 | 9.8 |
| Day 20 | 1277.6 | 374.9 | 47.2 | 29.9 |
| Day 25 | 3563.7 | 1045.8 | 113.5 | 72.0 |
| Day 30 | 7778.9 | 2282.9 | 215.8 | 136.9 |
| Day 35 | 13287.9 | 3899.7 | 324.6 | 206.0 |
| Day 40 | 17762.9 | 5213.0 | 386.2 | 245.0 |
| Day 45 | 18582.2 | 5453.5 | 363.4 | 230.6 |
| Log values were re-transformed to levels using the transformation method as described in Cameron & Trivedi<sup>23</sup>. The expected values were computed holding all other covariates at the sample mean. |  |  |  |  |

A higher percentage of African American population in the state was associated with significantly higher cumulative positive cases (β: 0.08, T-stat: 4.01). The results hold when the first-day fixed effects were applied to the model (β: 0.07, T-stat: 3.03). Using daily incremental positive cases as outcome also yielded significant results, both without (β: 0.06, T-stat: 3.50) and with FDFE (β: 0.06, T-stat: 3.24). These results were robust across all our sensitivity analyses. This implies that, when the share of African American population is higher by 1 percentage point, on average cumulative and daily rates were higher by 8 percent and 6 percent respectively. The percentage of state population in poverty showed a negative association with cumulative positive cases in the model without FDFE, but ceased to be significant in model with FDFE, and showed no significant association with daily incremental cases. This pattern persists in the sensitivity analysis.

We found strong evidence of a quadratic trend – with both cumulative and daily rates increasing at a decreasing rate. For example, for cumulative rates, the coefficient estimate of ‘Days’ was positive and significant (β: 0.43, T-stat: 22.04), and for Days^2^ was negative and significant (β: −0.01, T-stat: −20.68). With respect to the other covariates, we found that total state population was significantly associated with higher cumulative positive rates, though the relationship was not statistically significant for daily positive rates. Asthma prevalence was significantly associated with cumulative positive cases without FDFE, but no longer significant when FDFE was included. Nor did we see any consistent significant associations between cumulative or daily rates and population density, diabetes, and age above 65.

Our results did not change when we further included covariates of certified nursing facilities or physicians per 100,000 for states, nor were those covariates significant. We explored adding state prevalence of hypertension and obesity, but those variables were multicollinear with diabetes – with variance inflation factors of 4 or higher – and did not yield meaningful results.

## Discussion

We present one of the first analyses of state stay-at-home orders, state African American population and other state-level socio-demographic covariates on COVID-19 incidence rates. Our results provide support for the hypothesis that stay-at-home orders do have an impact on reducing COVID-19 infection rates. Expected predicted values from our main model suggest cumulative rates would be almost three times higher if stay-at-home orders had not been in place. Our results also strongly suggest that greater African American population is associated with higher COVID-19 infection rates, suggesting that African-Americans may be particularly vulnerable to infections. However, state poverty or prevalence of diabetes were largely insignificant. We did see a significant association between cumulative cases and asthma in our cumulative case main model. However, this result did not hold true for our other models. We did not see any association of rates with population density or population age above 65 years, though total population was significant in some models.

Unsurprisingly, there was a strong association between positive cases identified and the total number of tests administered. A substantial percentage of those infected are asymptomatic^25^. Hence, continued testing of symptomatic individuals supplemented by random testing will be essential to get accurate statistics on incidence of the disease and policy guidance and decisions going forward^26^.

Our study found evidence that stay-at-home orders were associated with substantially reducing the cumulative case rate and the daily case rate. Our findings correspond to earlier studies from Wuhan, China ^7^, and to county-level analyses in U.S. indicate that shutdowns could save lives^7, 27^, a four-state analysis indicating stay-at-home slowed the cumulative increase in hospitalizations^6^, and an Iowa-Illinois comparison indicating stay-at-homes orders reduced infection rates^5^. Thus, we add to the small but growing body of evidence that the shutdowns -- which were widely supported by infectious disease and public health experts -- have been helpful in curbing the disease. While long-term continuation of shutdowns are not feasible, lessons from the 1919 pandemic emphasize the importance of convincing the public about the need for restrictions via scientifically accurate and convincing messaging^28^. We hope that this study’s findings will strengthen the credibility of safety measures that health experts are continuing to recommend, like wearing masks and maintain social distancing.

Our study found strong associations between the percentage of African Americans in the state and the case rate, and this association persisted across all model specifications and sensitivity analyses. This supports existing reports from multiple sources^9^. While racial disparities in health in the U.S. are well established^29^, what was perhaps surprising is that factors like poverty or diabetes prevalence – speculated as being potential mediators between minority race and COVID-19 infections – were either not significant in our models, or had a counterintuitive negative sign. Hence, racial disparities in COVID-19 infections may have reasons beyond disparities in poverty or obesogenic conditions – for example, housing conditions, work circumstances or over-representation in prisons and detention centers can help explain this disparity^30^. Further investigation into this is important, since disparity figures by themselves “can perpetuate harmful myths and misunderstandings that actually undermine the goal of eliminating health inequities.” ^31^ We believe our findings are especially pertinent given the country-wide protests against police brutality on African Americans. It is a particularly poignant fact that the protests may expose an already vulnerable demographic group to even further risk of COVID-19 infections^32^, and more than 1000 scientists and community stakeholders have written an open letter to state and local governments on how to reduce public health risk during protests, while emphasizing the importance of supporting peaceful protests^33^.

Some of our findings are unexpected – for example, the statistical significance of the total state population coupled with the relative lack of significance of population density. This may be continued testimony to how little we know about predictors and risk factors for this disease, and that what ‘seems’ logical may not necessarily hold up under empirical scrutiny. We tentatively suggest that sporadic super-spreader events in low population-density states, for example a funeral in Georgia, or infection clusters in rural meat-packing facilities, may dilute the overall relationship between state population-density and case rates^34^.

### Limitations

Our study has several limitations. Its observational nature prevents making causal inferences. Additional limitations are as follows: First, testing protocols and testing availability is not uniform across states, and some tests may be unreported; further, there may be delays in reporting tests and positive cases, leading to inaccuracies in the daily data. Second, identified positive cases may substantially underestimate the actual number of positive cases. Third, the rigor with which ‘stay-at-home’ was enforced and adhered to may have varied from state to state, and also varied across areas within states, which is not captured in our data. Fourth, we could not account for local stay-at-home ordinances at the city or county level, since we lacked exhaustive information on such ordinances or what fraction of the state’s population was impacted by them. Fifth, our study used state-level socio-demographic characteristics, which may differ from the impact of those socio-demographic characteristics at a more granular level -– for example, the insignificant or negative relationship between poverty and COVID-19 that we see may not hold for zip-code or neighborhood-level analyses. Finally, we did not explore how trends may have changed following the state re-openings, though we intend to do this in forthcoming analyses.

In conclusion, we are rapidly moving into a phase where other more moderate measures will be replacing stay-at-home – which has been compared to shifting from ‘abstinence only’ to teaching safe-sex practices during the HIV epidemic^35^. The ongoing mass protests against police-brutality has added a further dimension to infection-risk which may also disproportionately impact African-Americans. Thus, it is imperative that we have continuing research on what factors curb COVID-19 contagion, as well as what factors underlie the vulnerability of minority races to this disease. Such research is essential to guiding decision-making, and helping reduce racial disparities in COVID-19 outcomes – which should be seen as part of the overarching goal of enhancing social justice and improving equity in the U.S.

## Data Availability

All data sources listed below

https://www.kff.org/other/state-indicator/distribution-by-raceethnicity/?currentTimeframe=0&sortModel=%7B%22colId%22:%22Location%22,%22sort%22:%22asc%22%7D.

https://docs.google.com/spreadsheets/u/2/d/e/2PACX-1vRwAqp96T9sYYq2-i7Tj0pvTf6XVHjDSMIKBdZHXiCGGdNC0ypEU9NbngS8mxea55JuCFuua1MUeOj5/pubhtml#

https://coronavirus.1point3acres.com/en

https://www.census.gov/data/tables/time-series/demo/popest/2010s-national-total.html

https://worldpopulationreview.com/states/state-densities/

https://www.census.gov/data/tables/2019/demo/income-poverty/p60-266.html

https://www.cdc.gov/brfss/data_documentation/index.htm

https://www.kff.org/other/state-indicator/total-active-physicians/?currentTimeframe=0&sortModel=%7B%22colId%22:%22Location%22,%22sort%22:%22asc%22%7D

https://www.kff.org/other/state-indicator/number-of-nursing-facilities/?currentTimeframe=0&sortModel=%7B%22colId%22:%22Location%22,%22sort%22:%22asc%22%7D

